# Comprehensive characterization of COVID-19 patients with repeatedly positive SARS-CoV-2 tests using a large US electronic health record database

**DOI:** 10.1101/2021.06.10.21256915

**Authors:** Xiao Dong, Yujia Zhou, Xiao-ou Shu, Elmer V. Bernstam, Rebecca Stern, David M. Aronoff, Hua Xu, Loren Lipworth

**Affiliations:** School of Biomedical Informatics, The University of Texas Health Science Center at Houston, Houston, TX; Division of Epidemiology, Department of Medicine, Vanderbilt University Medical Center, Nashville, TN; Division of General Internal Medicine, Department of Internal Medicine, McGovern Medical School, The University of Texas Health Science Center at Houston, Houston, TX; Division of Infectious Diseases, Department of Medicine, Vanderbilt University Medical Center, Nashville, TN

**Author notes:** Co-corresponding authors., Address correspondence to: Loren Lipworth, ScD, Division of Epidemiology, 2525 West End Avenue, Suite 300, Nashville, TN 37203, Hua Xu, PhD,7000 Fannin St, Suite 730E, Houston, TX 77025.

## Abstract

**Background:** In the absence of genome sequencing, two positive molecular SARS-CoV-2 tests separated by negative tests, prolonged time, and symptom resolution remain the best surrogate measure of possible re-infection.

**Methods:** Using a large electronic health record database, we characterized clinical and testing data for 23 patients with repeatedly positive SARS-CoV-2 PCR test results ≥60 days apart, separated by ≥2 consecutive negative test results. Prevalence of chronic medical conditions, symptoms and severe outcomes related to COVID-19 illness were ascertained.

**Results:** Median age was 64.5 years, 40% were Black, and 39% were female. 83% smoked within the prior year, 61% were overweight/obese, 83% had immune compromising conditions, and 96% had ≥2 comorbidities. Median interval between the two positive tests was 77 days. Among the 19 patients with 60-89 days between positive tests, 17 (89%) exhibited symptoms or clinical manifestations indicative of COVID-19 at the time of the second positive test and 14 (74%) were hospitalized at the second positive test. Of the four patients with ≥90 days between two positive tests, two had mild or no symptoms at the second positive test and one, an immune compromised patient, had a brief hospitalization at the first diagnosis, followed by ICU admission at the second diagnosis three months later.

**Conclusions:** Our study demonstrated a high prevalence of immune compromise, comorbidities, obesity and smoking among patients with repeatedly positive SARS-CoV-2 tests. Despite limitations, including lack of semi-quantitative estimates of viral load, these data may help prioritize suspected cases of reinfection for investigation and continued surveillance.

**Importance:** Comprehensive characterization of clinical and SARS-CoV-2 testing data for patients with repeatedly positive SARS-CoV-2 tests can help prioritize suspected cases of reinfection for investigation in the absence of sequencing data and for continued surveillance for potential long-term health consequences of SARS-CoV-2 infection.

## Introduction

As of 5 May 2021, there have been more than 155 million confirmed cases of COVID-19 globally, including 33 million in the United States. A reverse transcriptase polymerase chain reaction (RT-PCR) test is considered the gold standard for detection of SARS-CoV-2 in upper and lower respiratory specimens and for diagnosis of COVID-19. While neutralizing antibodies are detectable for several months following recovery from SARS-CoV-2 infection^1,2^, it remains unknown whether and for how long these antibody responses protect patients from re-infection. There have been many case reports of patients with a second positive PCR test after their PCR results turned negative and symptoms resolved^3^. Most of these are suspected cases of re-infection based on limited clinical or testing data; in a minority of suspected cases of reinfection, the viral genome sequences were analyzed and shown to be distinct, strongly supporting a re-infection rather than failure to clear an initial infection^4^. In the absence of genomic evaluations, the presence of two positive molecular tests separated by negative tests, prolonged time, and clinical resolution of symptoms remains the best surrogate measurement of possible re-infection. Using the Centers for Disease Control and Prevention Common Investigation Protocol for Investigating Suspected SARS-CoV-2 Reinfection^4^ as a guide, we conducted a comprehensive evaluation of patients who had repeated positive SARS-CoV-2 PCR tests in a large US COVID-19 electronic health record (EHR) database. We characterize their demographic and clinical characteristics, including their SARS-CoV-2 testing journey, symptoms, medication use and COVID-19 related complications.

## Patients and Methods

This retrospective study used the Optum® COVID-19 dataset^5^, which implements a low-latency data acquisition model that aggregates de-identified EHR data from providers across the continuum of care.

As of August 20, 2020, the Optum® COVID-19 dataset included 73,702 patients with a COVID-19 diagnosis code (ICD10 U07.1) that was laboratory-confirmed with a positive SARS-CoV-2 PCR test, of whom 690 had two positive PCR test results separated by at least one negative test result. The study sample was further restricted to patients who had two consecutive negative test results >24 hours apart between two positive test results (N=79); of these, 4 patients had at least 90 days between their two positive tests and another 19 had at least 60 days between their two positive tests and had accessible demographic and clinical data (Figure 1). If a negative test and a positive test were returned on the same day (<24h), both tests were disregarded.

**Figure 1.**
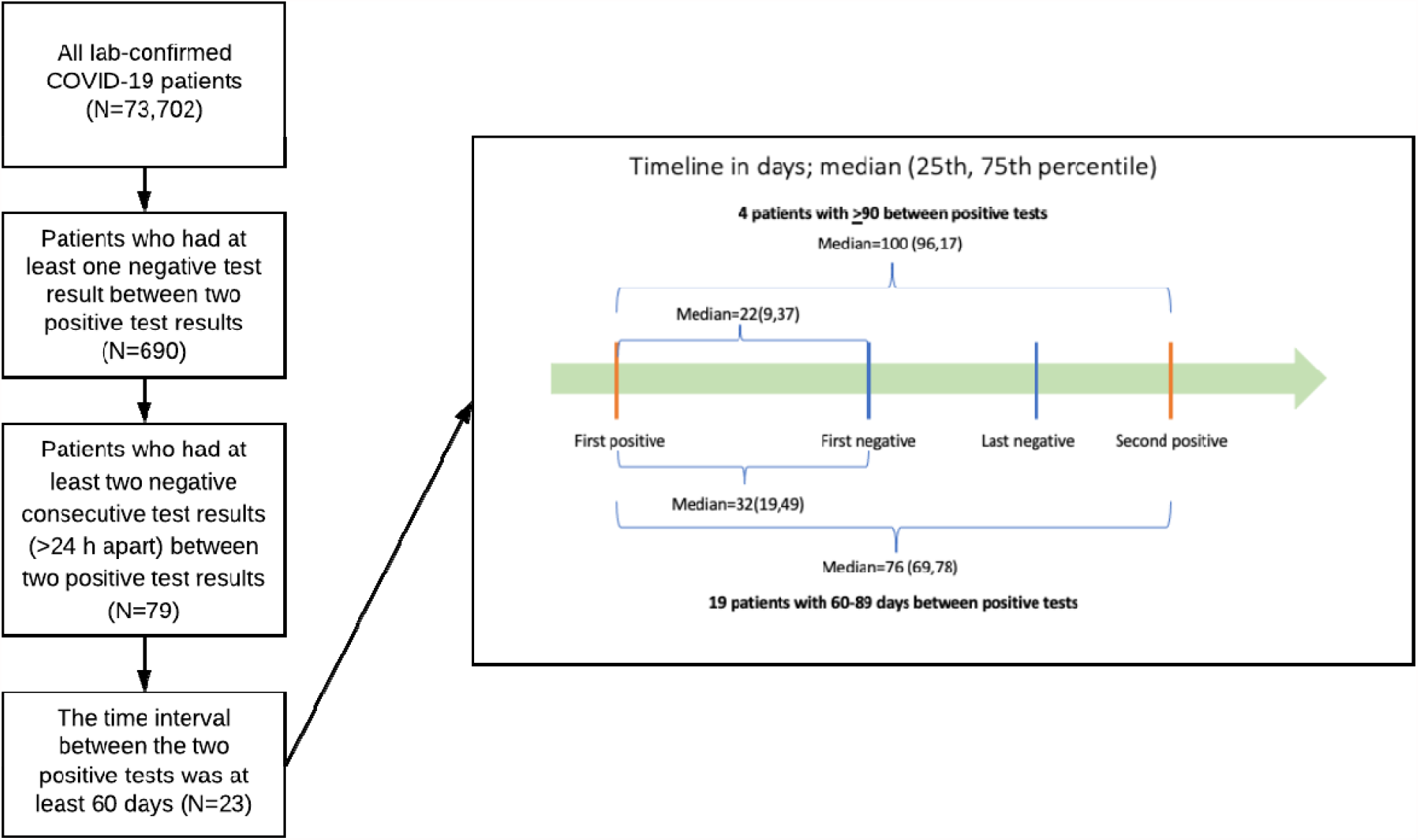
Study flow chart and SARS-CoV-2 PCR testing timeline (days) for 23 repeatedly positive patients.

Demographic and clinical information, including age, gender, race/ethnicity, smoking status, and body mass index (BMI), was extracted. Smoking and BMI were based on the patient’s most recent record within one year prior to the index date (first SARS-CoV-2 positive test date). Available EHR data for the 23 patients were manually reviewed. Prevalence of chronic medical conditions which are considered risk factors for COVID-19 was ascertained, including: insulin-dependent type 2 diabetes, hypertension, chronic kidney disease (CKD), respiratory disease (including chronic obstructive pulmonary disease), cardiovascular disease (CVD), atrial fibrillation and immune compromising conditions (including end-stage renal disease on dialysis, HIV, cirrhosis including alcohol-related, solid organ transplant, cancer, and protein-calorie malnutrition). Symptoms typical of COVID-19 were ascertained for each patient during each of two time periods, within 30 days before and after the index date and the second positive test date. In addition, severe clinical outcomes related to COVID-19 illness and medications commonly used to treat COVID-19 were ascertained during each time period, including the following: hospitalization, intensive care unit (ICU) admission, mechanical ventilation, tracheostomy, amputation, or death (at second positive test).

Continuous variables were expressed as median (25^th^, 75^th^ percentile), and categorical variables as counts (percentages). Missing data were not imputed.

## Results

For the four patients with at least 90 days between positive tests, the median interval between the two positive tests, separated by two or more consecutive negative tests 24 hours apart, was 100 days (25^th^, 75^th^ percentile: 96, 107), and the median interval between the first positive and first negative test was 22 days (9, 37) (Figure 1). For the 19 patients with 60-89 days between positive tests, the corresponding intervals were 76 days (25^th^, 75^th^ percentile: 69, 78) and 32 days (19, 49), respectively (Figure 1).

Median age of the 23 repeatedly positive patients at the index date was 64.5 years (25^th^, 75^th^: 53.5, 69.8). Seventeen patients were diagnosed in the Northeast, five in the Midwest and one in the South; 40% of patients were Black, 40% white, and 20% other/unknown race, 83% had non-Hispanic ethnicity, and 39% were female. Almost 83% smoked within the prior year, and 61% were overweight or obese.

Comorbidity diagnoses and symptom prevalence for the 23 individual patients at the time of each positive test are presented in Table 1, and their PCR test and clinical journeys are shown in Figure 2. Chronic disease prevalence was high, including hypertension (70%), CVD, atrial fibrillation or CKD (each 26%), and insulin-dependent type 2 diabetes or history of venous thromboembolism/long-term anticoagulation (each 22%). Overall, 96% of patients had ≥2 comorbidities. Most notably, 19 of the patients (83%) had immune compromising conditions, including two of the four patients with ≥90 days between positive tests (PT14 and PT19).

**Figure 2.**
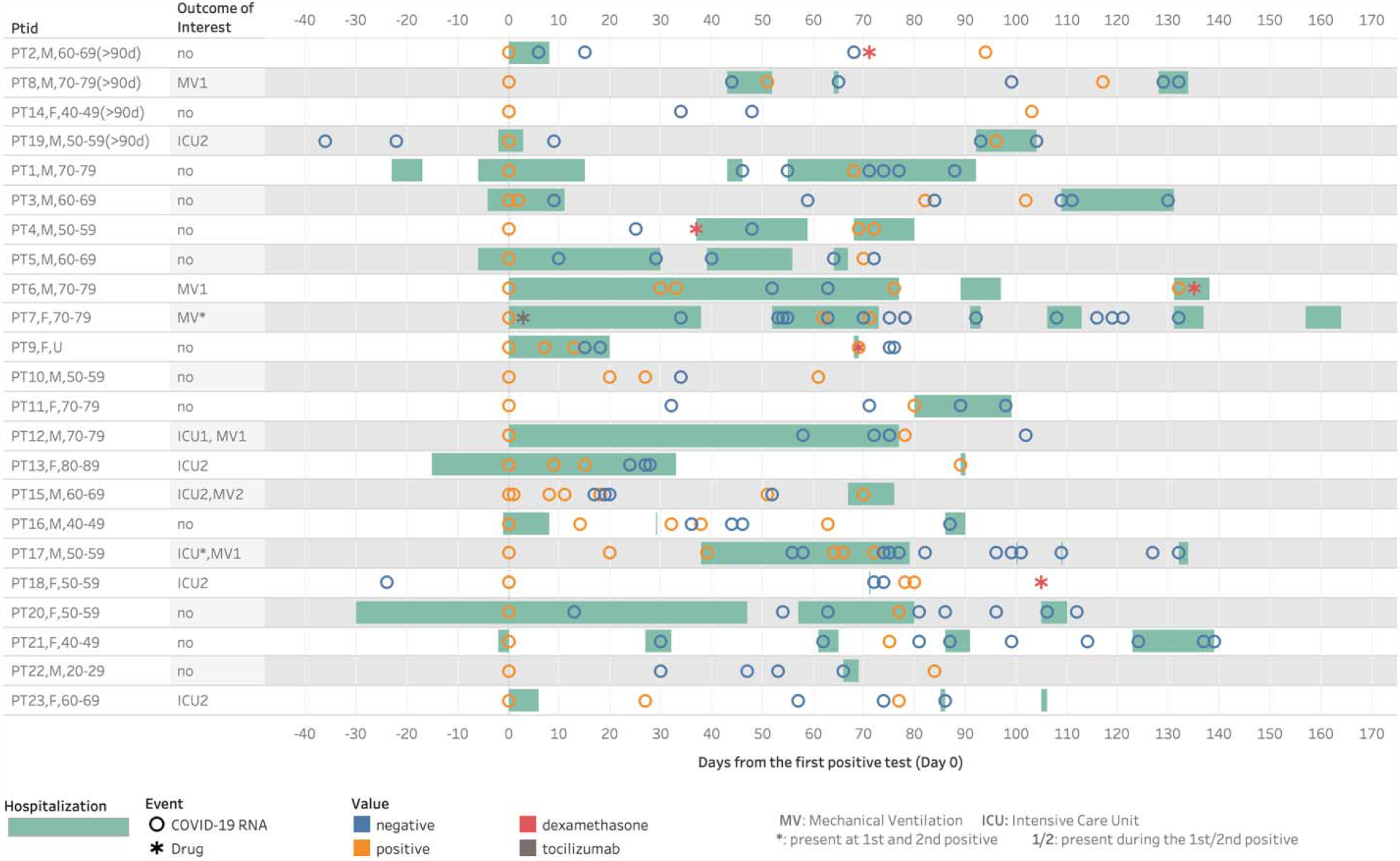
COVID-19 RT-PCR testing and clinical journey for 23 patients with repeatedly positive tests

For individuals with 45-89 days between positive SARS-CoV-2 tests, CDC investigative criteria include having “a symptomatic second episode and no obvious alternate etiology for COVID-19–like symptoms OR close contact with a person known to have laboratory-confirmed COVID-19.” Among the 19 patients in our study with 60-89 days between positive tests, 17 (89%) exhibited symptoms or clinical manifestations indicative of COVID-19 at the time of the second positive test, including 9 (47%) with acute respiratory failure, 8 (42%) with acute kidney failure, 6 (32%) with shortness of breath, 5 (26%) with fever, and 3 with acute embolism and thrombosis (16%). Fourteen of the 19 (74%) were hospitalized at the second positive test, all but four of whom were also hospitalized at the first positive COVID-19 test. One patient was treated with tocilizumab (PT7, at the time of first positive test and during an extended hospitalization) and 4 were treated with dexamethasone after the first diagnosis of COVID-19.

As shown in Figure 2 and Table 1, four of the patients (PT5, PT7, PT12, and PT17) with immune compromising conditions had severe symptoms and lengthy hospitalizations (including ICU and mechanical ventilation for PT12 and PT17) beginning at the first positive COVID-19 test and numerous negative tests, often with their second positive test in close proximity to one or multiple negative tests. Additionally, PT10 had no COVID-19-like symptoms or related treatments at the second positive test, and PT18 had esophageal cancer and no COVID-19-like symptoms at the time of either positive test. The clinical journeys of these six repeatedly positive patients cast doubt about the accuracy of categorizing them as true re-infections.

Of the four patients (Figure 2, top) who had ≥90 days between two positive tests, the record of one immune compromised patient (PT14) suggests mild-to-moderate disease with few symptoms following both COVID-19 diagnoses. PT19, also immune compromised, had a brief hospitalization at the first diagnosis, followed by ICU admission at the second diagnosis three months later. PT2 had severe symptoms and hospitalization and treatment with dexamethasone after the first positive test, but no symptoms or treatment at the second positive test.

No patients had cardiac arrest, tracheostomy, amputation, or death.

## Discussion

This study provides clinical and testing characterization of 23 COVID-19 patients with suspected re-infection, defined as repeatedly positive SARS-CoV-2 PCR tests separated by consecutive negative tests and prolonged time. We observed a high prevalence of Black race, obesity, and multiple comorbidities known to increase risk of COVID-19 illness, including hypertension, diabetes and CKD. Moreover, 83% of the patients with repeated positivity were current smokers, which is linked to increased risk of severe COVID-19^6^. It is possible that those known to be at particularly high risk for COVID-19 or those with persistent or recurrent symptoms may undergo frequent testing, thereby increasing the likelihood of receiving some false positive or false negative results.

Immune compromising conditions including end-stage renal disease on dialysis, HIV, cirrhosis including alcohol-related, solid organ transplant, cancer, and protein-calorie malnutrition were more common in our study population affected by COVID-19 compared to the general population without COVID-19. Among the subset of patients in our study with immune compromising conditions, more than two-thirds required hospitalization for the second positive PCR test after the interval negative PCR test. Reinfection may therefore raise clinical suspicion for an underlying immune defect, which may have also influenced duration to achieve viral clearance.

Recent studies focused on immune-compromised populations with COVID-19 have highlighted elevated risks of COVID-19 severity and morbidity, as well as frequent multimorbidity. High attributable 28-day mortality due to COVID-19 for patients in the ERA-EDTA Registry, including 3285 on dialysis and 1013 with a functional kidney transplant, was substantial at 20% or 21-times higher and 19.9% or 92-times higher, respectively, compared to matched controls^7^. Similar trends of increased mortality and worsening liver function tests have been demonstrated in patients with cirrhosis and COVID-19^8^.

Moreover, alcohol-related liver disease and baseline hepatic dysfunction have been shown to be independent risk factors for death related to COVID-19^9^. Patients with HIV in our sample had at least one other significant comorbidity, either alcoholic cirrhosis (PT4, PT14) or ESRD on HD (PT20). This aligns with findings of an observational prospective study in Madrid by Vizcarra *et al*. describing 51 HIV-positive patients with COVID-19 having a significantly higher prevalence of comorbidities and age-adjusted mortality^10^.

Over 90% of patients in our study exhibited symptoms or clinical manifestations indicative of COVID-19 at the time of the second positive test, thereby fulfilling an important CDC criterion for the investigation of suspected SARS-CoV-2 reinfection, particularly among those with 45-89 days between positive PCR tests. Over three-fourths were hospitalized at the second positive test. Overall, 70% (12/17) of patients hospitalized at the first positive test were also hospitalized at the second test, suggesting that in most cases re-infection was not associated with less severe disease. This proportion is somewhat inconsistent with the finding of a recent review of 16 reported cases of re-infection confirmed by sequencing^11^, in which the severity of the re-infection episode was asymptomatic/mild in 75% of cases. Overall, in our study, 37% of those hospitalized had severe disease characterized by ICU admission. This is higher than previous estimates that 17-35% of hospitalized COVID-19 patients are treated in an ICU^2^. Acute kidney injury (AKI) has been reported to occur in approximately 9% of hospitalized COVID-19 patients and a higher proportion of those requiring ICU admission^2^. We observed AKI as a more common complication associated with COVID-19, including after both COVID-19 diagnoses in several individuals, but we were unable to determine whether these were independent or persistent events.

It is possible that hospitalized patients are more likely to undergo frequent testing, due to more severe disease or to support discharge to a rehabilitation facility or nursing home. This frequent testing can lead to alternating positive and negative tests, often on overlapping days. Given the high hospitalization rate in our study, repeated positive tests for some patients (e.g., PT5, PT7, PT12 and PT17) occurring during the time of an extended hospitalization with severe complications, including need for ICU admission and/or mechanical ventilation, may not represent true re-infections. Moreover, prolonged viral shedding, as has been observed in severe COVID-19 cases^12^, cannot be ruled out. A recent analysis in the Emory Healthcare System indicated that, among 22,443 patients who had at least two tests, the median (IQR) duration between first and last positive test was 19 days (12, 32), and a duration of 45 and 90 days represented the 88^th^ and 97^th^ percentile, respectively^11^.

In the absence of genomic evaluations to definitively confirm reinfection^3,11^, finding two positive molecular tests separated by negative tests, prolonged time, and resolution of symptoms remains the best surrogate measure of possible re-infection. In 70 previously reported cases to date^3^, with an average of 101 days between first and second positive test, viral genome sequences were shown to be distinct, strongly suggesting a re-infection rather than failure to clear an initial infection. Our identification and clinical characterization of 23 possible re-infections in a large dataset, with a median of 77 days between positive tests, provides additional data suggesting that re-infections may be common. Since most patients in the Optum dataset did not have repeated tests after their COVID-19 diagnosis, the true incidence rate of recurrent detectable SARS-CoV-2 cannot be estimated.

Our analysis was limited by lack of information on RT-PCR platforms (with varying sensitivities) or semi-quantitative RT-PCR cycle threshold (Ct) values. The patients in our study nevertheless fulfilled CDC criteria for cases ≥90 days apart or 45-89 days apart based on positive RT-PCR, and based on our study definition, cases were classified as re-infection rather than relapse based on interval negative RT-PCR. We were not able to confirm if COVID-19 was the primary diagnosis prompting hospitalization, if patients were incidentally found to test positive for SARS-CoV-2 upon admission for an unrelated illness, or later became symptomatic during the hospitalization course. Diagnoses may be more likely to be incidental if associated with ICD-10 codes for pre-procedural exam (e.g., elective surgery); however, a pre-procedural exam at the time of the second positive test was noted for only one patient in our study (PT2). It is noteworthy that among the 23 patients with confirmed RT-PCR re-positivity for SARS-CoV-2, a minority were not assigned an ICD-10 code for COVID-19 in the EHR. This did not appear to be associated with severity of disease presentation. Finally, repeatedly positive tests do not necessarily mean a re-infection, and persistent infection or relapse cannot be ruled out, particularly if signs and symptoms observed at the second positive test are similar to those seen in individuals with post-acute sequelae of COVID-19 (or “long COVID”).

Despite these limitations, our study provides a comprehensive characterization of demographic, clinical and SARS-CoV-2 testing data for patients with repeatedly positive SARS-CoV-2 tests in a large EHR database across the US, which could help prioritize suspected cases of reinfection for investigation in the absence of sequencing data and for continued surveillance for potential long-term health consequences of SARS-CoV-2 infection. Further investigation into risk of reinfection by type and degree of immunosuppressive condition, medications, and disease chronicity will be valuable for future goals of prevention, mitigation of risk factors, and reducing severity of illness.

## Data Availability

Optum's COVID-19 data are available for researchers for approved research purposes. By negotiation with Optum, the dataset is potentially accessible to any qualified investigator within the University of Texas System. It can be used in conjunction with grants, but as required by Optum, not for pharmaceutical industry-funded projects.
University of Texas Health Science Center-Houston. Center for Healthcare Data (CHCD). https://dataportal.uta.edu/node/27. Published 2021. Accessed May 6, 2021

## References

1. Wajnberg A, Amanat F, Firpo A, et al. Robust neutralizing antibodies to SARS-CoV-2 infection persist for months. Science. 2020;370(6521):1227–1230.

2. Wiersinga WJ, Rhodes A, Cheng AC, Peacock SJ, Prescott HC. Pathophysiology, Transmission, Diagnosis, and Treatment of Coronavirus Disease 2019 (COVID-19): A Review. JAMA. 2020;324(8):782–793.

3. BNO News. COVID-19 Reinfection Tracker. https://bnonews.com/index.php/2020/08/covid-19-reinfection-tracker/. Accessed May 4, 2021.

4. Centers for Disease Control and Prevention. Common Investigation Protocol for Investigating Suspected SARS-CoV-2 Reinfection. https://www.cdc.gov/coronavirus/2019-ncov/php/reinfection.html. Published 2020. Accessed May 4, 2021.

5. University of Texas Health Science Center-Houston. Center for Healthcare Data (CHCD). https://dataportal.uta.edu/node/27. Published 2021. Accessed May 6, 2021.

6. Reddy RK, Charles WN, Sklavounos A, Dutt A, Seed PT, Khajuria A. The effect of smoking on COVID-19 severity: A systematic review and meta-analysis. J Med Virol. 2020.

7. Jager KJ, Kramer A, Chesnaye NC, et al. Results from the ERA-EDTA Registry indicate a high mortality due to COVID-19 in dialysis patients and kidney transplant recipients across Europe. Kidney Int. 2020;98(6):1540–1548.

8. Iavarone M, D’Ambrosio R, Soria A, et al. High rates of 30-day mortality in patients with cirrhosis and COVID-19. J Hepatol. 2020;73(5):1063–1071.

9. Marjot T, Moon AM, Cook JA, et al. Outcomes following SARS-CoV-2 infection in patients with chronic liver disease: An international registry study. J Hepatol. 2021;74(3):567–577.

10. Vizcarra P, Pérez-Elías MJ, Quereda C, et al. Description of COVID-19 in HIV-infected individuals: a single-centre, prospective cohort. Lancet HIV. 2020;7(8):e554–e564.

11. Babiker A, Marvil CE, Waggoner JJ, Collins MH, Piantadosi A. The Importance and Challenges of Identifying SARS-CoV-2 Reinfections. J Clin Microbiol. 2021;59(4).

12. Cevik M, Tate M, Lloyd O, Maraolo AE, Schafers J, Ho A. SARS-CoV-2, SARS-CoV, and MERS-CoV viral load dynamics, duration of viral shedding, and infectiousness: a systematic review and meta-analysis. Lancet Microbe. 2021;2(1):e13–e22.

