## Supplementary material for "Comprehensive characterization of COVID-19 patients with repeatedly positive SARS-CoV-2 tests using a large US electronic health record database": Table 1

| **Table 1**. Individual-level comorbidity and symptom data for the 23 repeatedly positive patients, based on manual review of ICD-10 codes within 30 days before and after the index date and the second positive test date | | | | | | | | |
| --- | --- | --- | --- | --- | --- | --- | --- | --- |
|  | **≥90 days between positive SARS-CoV-2 tests** | | | | **60-89 days between positive SARS-CoV-2 tests** | | | |
| Patient | 2 | 8 | 14 | 19 | 1 | 3 | 4 | 5 |
| Co-morbidities | HTN  HLD | Nicotine  AFib  HTN  Pacemaker  Long QT  h/o VTE  Long-term AC | Nicotine  HIV  Alcoholic cirrhosis  Protein-calorie malnutrition  Alcohol dependence  TB | HTN  Alcohol dependence | Insulin-dependent DM2 w/CKD  COPD  Nicotine  AFib  HTN  HLD  NSTEMI  Pacemaker  Long QT  h/o VTE  Long-term AC  ESRD on HD  Cancer - retroperitoneum | Kidney-heart transplant  Rheumatoid arthritis  Protein-calorie malnutrition | HTN  HLD  HIV  Alcoholic cirrhosis w/ascites | Prostate cancer  Alcoholic cirrhosis w/ascites  Alcohol abuse  h/o Pulmonary TB |
| Symptoms COVID-19 episode #1 | SOB, cough, fever, chest pain, pneumonia, acute respiratory failure w/hypoxia, bradycardia | NONE | Low back pain | Fever, pneumonia, hypoxia, AKI | SOB, diarrhea, weakness, low back pain, pneumonia, acute respiratory failure w/hypoxia, ARDS, altered mental status, metabolic encephalopathy, fluid overload, ventricular tachycardia | Cough, chest pain, pneumonia, AKI, tachycardia | Fever, tachycardia | Cough, fever, headache, chest pain, tachycardia, acute embolism and thrombosis |
| Symptoms COVID-19 episode #2 | NONE | SOB, bradycardia | Fever | fever, respiratory failure w/hypoxia, encephalopathy, AKI | acute respiratory failure w/hypoxia, ventricular tachycardia | SOB, diarrhea, respiratory failure w/hypoxia, AKI, tachycardia | SOB, fever, AKI, tachycardia, acute embolism and thrombosis | Chest pain, tachycardia, acute embolism and thrombosis |

Continued

| Patient | 6 | 7 | 9 | 10 | 11 | 12 | 13 | 15 |
| --- | --- | --- | --- | --- | --- | --- | --- | --- |
| Co-morbidities | HTN  HLD  Old MI  NSTEMI  Long QT  ESRD on HD  Protein-calorie malnutrition  OSA  Thyroid cancer | Insulin-dependent DM2 w/CKD  AFib  HTN  HLD  Long QT  Long-term AC  CKD  Kidney transplant  Protein-calorie malnutrition | AFib  HTN  HLD  Old MI  Pacemaker  Long QT  Long-term AC  CKD  Breast cancer  Protein-calorie malnutrition | HTN  CLL w/o remission | Insulin-dependent DM2  Nicotine  HTN  HLD  Old MI  Cirrhosis | AFib  HTN  HLD  CKD  Protein-calorie malnutrition | AFib  HTN  HLD  Long-term AC  Cirrhosis  Protein-calorie malnutrition | Insulin-dependent DM2 w/CKD  HTN  HLD  Old MI  ESRD on HD  Heart transplant |
| Symptoms COVID-19 episode #1 | Fever, diarrhea, pneumonia, acute respiratory failure w/hypoxia, ARDS, ventilator dependence, AKI, encephalopathy, tachycardia, severe sepsis w/shock, acute embolism and thrombosis | SOB, cough, headache, pneumonia, acute respiratory failure with hypoxia, ARDS, AKI, encephalopathy, fluid overload, sepsis w/shock, acute embolism and thrombosis | Weakness, pneumonia, acute respiratory failure w/hypoxia, AKI | SOB, fever, headache | Altered mental status | SOB, diarrhea, pneumonia, acute respiratory failure w/hypoxia, ARDS, AKI, encephalopathy, tachycardia, sepsis w/shock | pneumonia, acute respiratory failure w/hypoxia, tachycardia | Fever, headache, diarrhea, weakness, low back pain, pneumonia, acute respiratory failure w/hypoxia, metabolic encephalopathy, fluid overload |
| Symptoms COVID-19 episode #2 | Diarrhea, pneumonia, acute respiratory failure w/hypoxia, ARDS, AKI, tachycardia, sepsis w/shock, acute embolism and thrombosis | Fever, pneumonia, acute respiratory failure w/hypoxia, ventilator dependence, AKI, fluid overload, sepsis w/shock | Diarrhea, weakness, pneumonia, acute respiratory failure w/hypoxia, AKI | NONE | Weakness, AKI, metabolic encephalopathy, altered mental status, bradycardia | SOB, diarrhea, chest pain, pneumonia, acute respiratory failure w/hypoxia, ARDS, AKI, encephalopathy, tachycardia, sepsis w/shock | Diarrhea, chest pain, pneumonia, acute respiratory failure w/hypoxia, AKI, encephalopathy, sepsis w/shock | Weakness, pneumonia, acute respiratory failure w/hypoxia, encephalopathy, fluid overload, tachycardia, sepsis w/o shock |

Continued

| Patient | 16 | 17 | 18 | 20 | 21 | 22 | 23 |
| --- | --- | --- | --- | --- | --- | --- | --- |
| Co-morbidities | HTN  HLD | HTN  Protein-calorie malnutrition | Esophageal cancer | Insulin-dependent DM2 w/CKD  Nicotine  HTN  ESRD on HD  HIV | Nicotine  Long QT | Monoclonal gammopathy  Respiratory TB  Histoplasmosis  Blastomycosis | COPD  Nicotine |
| Symptoms COVID-19 episode #1 | Weakness, pneumonia, AKI, metabolic encephalopathy | Pneumonia, acute respiratory failure w/hypoxia, AKI, encephalopathy | NONE | Fever, headache, chest pain, acute respiratory failure w/hypoxia | Low back pain | NONE | Pneumonia |
| Symptoms COVID-19 episode #2 | Fever, weakness, chest pain, pneumonia, tachycardia | SOB, pneumonia | NONE | Chest pain | Low back pain | Fever | SOB, pneumonia, acute respiratory failure w/hypoxia |

*immune compromising condition

Abbreviations:

MI = myocardial infarction; AC = anticoagulation; HTN = hypertension; HDL = hyperlipidemia; SOB = shortness of breath; DM2 = type 2 diabetes mellitus; CKD = chronic kidney disease; AFib = atrial fibrillation; VTE = venous thromboembolism; ESRD = end-stage renal disease; HD = hemodialysis; TB = tuberculosis; OSA = obstructive sleep apnea; AKI = acute kidney injury; ARDS = acute respiratory distress syndrome; CLL = chronic lymphocytic leukemia
